# Ochratoxin A status at birth is associated with reduced birthweight and ponderal index in rural Burkina Faso

**DOI:** 10.1101/2024.04.19.24306069

**Authors:** Yuri Bastos-Moreira, Alemayehu Argaw, Giulianmichela Di Palma, Trenton Dailey-Chwalibóg, Jasmin El-Hafi, Lionel Olivier Ouédraogo, Laeticia Celine Toe, Sarah De Saeger, Carl Lachat, Marthe De Boevre

**Affiliations:** Center of Excellence in Mycotoxicology and Public Health, MYTOX-SOUTH® Coordination Unit, Faculty of Pharmaceutical Sciences, Ghent University, Ghent, Belgium; Department of Food Technology, Safety and Health, Faculty of Bioscience Engineering, Ghent University, Ghent, Belgium; Department of Chemistry, University of Torino, Torino, Italy; Institute of Food Chemistry, University of Münster, Münster, Germany; Laboratoire de Biologie Clinique, Centre Muraz, Bobo-Dioulasso, Burkina Faso; Unité Nutrition et Maladies Métaboliques, Institut de Recherche en Sciences de la Santé (IRSS), Bobo-Dioulasso, Burkina Faso; Department of Biotechnology and Food Technology, Faculty of Science, University of Johannesburg, Doornfontein Campus, Gauteng, South Africa

**Keywords:** birth outcomes, exposomics, growth, low- and middle-income countries, MISAME-III, mycotoxins, sub-Saharan Africa, ochratoxin A

## Abstract

**Background:** Mycotoxin exposure during pregnancy has been associated with adverse birth outcomes and poor infant growth. We assessed multiple biomarkers and metabolites of exposure to mycotoxins at birth and their associations with birth outcomes and infant growth in 274 newborns in rural Burkina Faso.

**Methods and findings:** Whole blood microsamples were analyzed for mycotoxin concentrations in newborns in the Biospecimen sub-study nested in MISAME-III trial using ultra performance liquid chromatography coupled to tandem mass spectrometry. Unadjusted and adjusted associations between mycotoxin exposure, and birth outcomes and infant growth at 6 months were estimated using linear regression models for continuous outcomes and linear probability models with robust variance estimation for binary outcomes. Infant growth trajectories from birth to 6 months were compared by exposure status using mixed-effects models with random intercept for the individual infant and random slope for the infant’s age. Ochratoxin A (OTA) exposure was detected in 38.3% of newborns, with other mycotoxins being detected in the range of 0.36% and 4.01%. OTA exposure was significantly associated with adverse birth outcomes, such as lower birthweight (β (95% CI): −0.11 kg (−0.21, 0.00); *p* = 0.042) and ponderal index (β (95% CI): −0.62 gm/cm^3^ (−1.19, −0.05); *p* = 0.034), and a marginally significant lower height growth trajectories during the first 6 months (β (95% CI): −0.08 cm/mo (−0.15, 0.0); *p* = 0.057).

**Conclusions:** OTA exposure was prevalent among newborns and also associated with lower growth at birth and during the first 6 months. The results emphasize the importance of nutrition-sensitive strategies to mitigate dietary OTA, as well as adopting food safety measures in Burkina Faso during the fetal period of development.

## Background

Mycotoxins are toxic fungal secondary metabolites that contaminate a wide spectrum of essential foods worldwide, including staple crops consumed by the most vulnerable populations (1). Foodstuffs in West Africa are commonly affected by mycotoxins (2,3) since the climate, where there is high temperature and humidity, is favorable for their production (4,5). Maternal nutrition affects both the pregnancy’s process and the newborn’s well-being (6). In low-and middle-income countries (LMICs), adverse pregnancy outcomes are common including low birth weight (LBW), preterm birth (PTB) and/or small-for-gestational age (SGA) (7). Several epidemiological studies have indicated that mycotoxin exposure is extensive in newborns (8–11).

The International Agency for Research on Cancer (IARC) categorizes aflatoxin B1 (AFB1) as carcinogenic to humans (Group1), and fumonisin B1 (FB1) and ochratoxin A (OTA) as possible human carcinogens (Group 2B) (12). The human fetus is vulnerable to health effects resulting from *in utero* exposure to environmental chemicals (13). Formerly, higher AF exposure, *in utero* and in early life, has been linked with stunting and/or underweight, while children with high fumonisins exposure were also shorter and lighter (14). In addition, research has also shown that OTA can cross the placental barrier in humans (15), and is reported to also have other toxic effects in humans including immunotoxicity and nephrotoxicity (16–18).

A literature review by Arce-López *et al*. (2020) concluded that OTA is often detected in whole blood, plasma and serum samples (19). Authors reported frequency levels of 64.9% (20–23), and concluded that the global population is generally exposed to OTA due to its long half-life in these matrices (19). This exposure during the critical first 1,000 days of life (10) might contribute to adverse fetal and infant outcomes (24). Generally, birthweight is an indicator of both maternal health and nutrition status, and also the infant’s well-being. Infants born with LBW are at increased risk of several short- and long-term consequences, including neonatal mortality, childhood stunting and impaired immune function (25–27). Nevertheless, research investigating the association between mycotoxins exposure and birth and infant growth outcomes have reported inconsistent results (28–31).

In Burkina Faso, limited biological and toxicological food contamination data are available (32), and legislation and regulations regarding mycotoxins are often not implemented (33,34). Using data from the Biospecimen (BioSpé) sub-study of the MISAME-III (MIcronutriments pour la SAnté de la Mère et de l’Enfant) trial in rural Burkina Faso, we previously reported a prenatal exposure to multiple mycotoxins among pregnant women from a rural Burkinabé setting, and found no evidence of associations with adverse birth outcomes and infant growth (in publication (35)). In the present study, we aimed to quantify newborn mycotoxin exposure at birth and investigated the association with birth outcomes and infant growth in the same mother-newborn dyads.

## Methods

### Study setting, participants, and design

Study protocols for the main MISAME-III trial (36) and the BioSpé sub-study nested under the MISAME-III trial (37) were published previously. The main MISAME-III study is a 2 x 2 factorial randomized controlled trial evaluating the effect of balanced energy-protein (BEP) supplementation to mothers during pregnancy (prenatal intervention) and lactation (postnatal intervention) on maternal and child outcomes. In a subsample from the main MISAME-III trial (Figure 1), a BioSpé sub-study was conducted aiming to understand the physiologic mechanisms through which the BEP supplement affects the maternal and child outcomes by way of multi-omics analyses, human biomonitoring of contaminants (mycotoxins, black carbon, gut enteropathogens and pesticides), and analysis of relative telomere length and mitochondrial DNA content (37).

**Figure 1.**
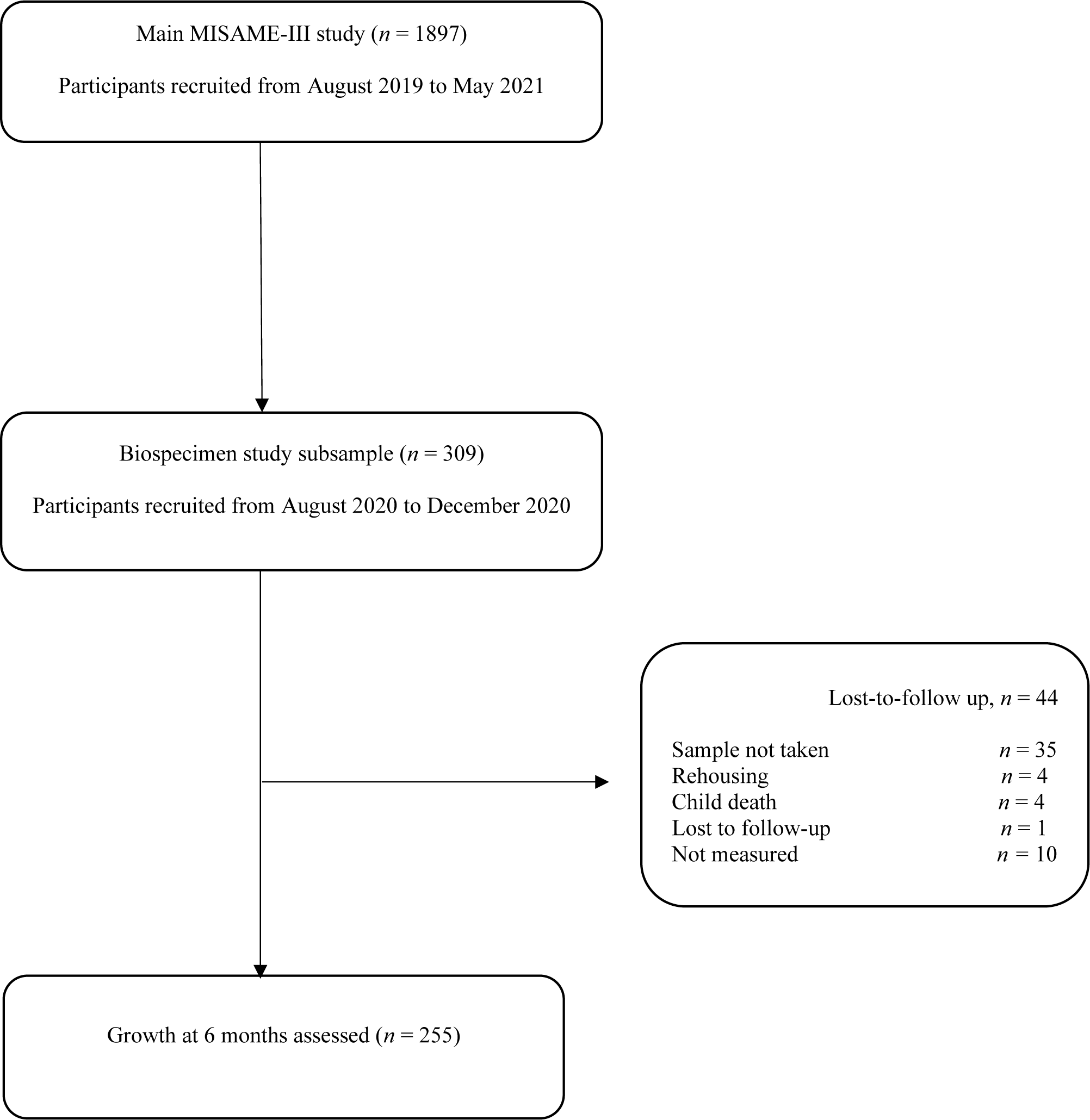
Study flow diagram of the Biospecimen sub-study (BioSpé) of the MISAME-III project.

The study was conducted in 6 rural health center catchment areas in the district of Houndé in the Hauts-Bassins region of Burkina Faso. The study area is characterized by a Sudano-Sahelian climate with a dry season running between September/October and April, and agricultural activities being the main livelihood of the community. Results from a previously conducted dietary survey in a sub-sample of the MISAME-III pregnant women showed the habitual diet during pregnancy is nondiverse, predominantly based on maize with a complement of leafy vegetables (38). Grains, roots, tubers and plantains together contributed 68% of the total calorie intake during pregnancy. Almost all participants (95%) consumed the main staple dish tô, which contributed 42% of the total energy intake. Tô is a stiff maize dough often served with a watery sauce containing green-leafy vegetables (okra, hibiscus, and baobab leaves) or other vegetables such as eggplant, with or without meat, fish, or caterpillars. Other food groups such as fruits, dairy, eggs, fish, and meat contributed very small amounts to the total energy intake (39).

### Exposure and outcomes

The present study considered exposure to a range of mycotoxins listed in Table 2. However, aflatoxin B1 (AFB1)-lysine exposure was not assessed due to the current unavailability of the commercial analytical standard for this specific mycotoxin. Mycotoxin exposure was defined as the detection of a concentration ≥ the limit of detection (LOD) in whole blood microsamples.

The outcomes of interest were birth outcomes, such as birth weight, SGA, LBW, gestational age (GA), PTB, length, mid-upper arm circumference (MUAC), head circumference, ponderal index, chest circumference, and infant growth and nutritional status at the age of 6 months, such as length-for-age z-score (LAZ), weight-for-age z-score (WAZ), weight-for-length *z*-score (WLZ), MUAC, head circumference, hemoglobin, stunting, underweight, wasting and anemia. We additionally assessed the associations between mycotoxin exposure and infant growth trajectories (height, weight, upper-arm and head circumferences) during the first 6 months postpartum.

PTB was defined as the birth of a newborn before 37 completed weeks of gestation. SGA was defined as a newborn weight less than the 10^th^ percentile of weight for the same GA and sex according to the International Fetal and Newborn Growth Consortium for the 21^st^ Century (INTERGROWTH-21^st^) (40). Anthropometric *z*-scores of LAZ, WAZ and WLZ were calculated based on the WHO Child Growth Standards with stunting, underweight and wasting defined as LAZ, WAZ and WLZ values below 2 SD from the median value for same age and sex from the reference population (41). Newborn Rohrer’s ponderal index was calculated as weight in g divided for length in cm cubed (i.e., weight/length^3^ (g/cm^3^) × 1,000).

### Data collection

The MISAME-III trial data were collected through computer-assisted personal interviewing using SurveySolutions (version 21.5) on tablets and then transferred to a central server at Ghent University. Sociodemographic and other relevant characteristics of participants and study households were collected at baseline during the first and early second trimester of pregnancy. All newborn anthropometry measurements were taken within 12 hours of birth, whereas mothers were invited for follow-up growth assessment every month until 6 months of age. Measurements were taken in duplicates and a third measurement was taken in case of a large discrepancy between the duplicate measurements. Length was measured to the nearest 1 mm with a Seca 416 Infantometer, weight was measured to the nearest 10 g with a Seca 384 scale, and head circumference, thoracic circumference and MUAC were measured to the nearest 1 mm with a Seca 212 measuring tape. GA was determined using a portable ultrasound (SonoSite M-Turbo, FUJIFILM SonoSite, Bothell, Washington, USA) during the first and early second trimester of pregnancy.

### Blood sample collection and laboratory analysis

Samples collection and lab analysis procedures were described in detail previously (37). Newborn samples were collected between May and October 2021 within 12 hours of birth in all newborns. An amount of 40 of capillary whole blood was collected by capillary sampling onto VAMS tips (2 × 20 µL VAMS tips), namely Mitra^TM^, via direct heel incision for mycotoxins analysis (37). Then, VAMS tips were stored in 20 µL Mitra Clamshells and transported from the health centers to the Institut de Recherche en Sciences de la Santé in Bobo-Dioulasso, Burkina Faso for shipment at room temperature to the Centre of Excellence in Mycotoxicology and Public Health, Faculty of Pharmaceutical Sciences, Ghent University, Belgium. For storage at −80°C until analysis, VAMS were placed in Mitra Autoracks (96-Sampler, item number: 108) inside a storage bag containing desiccant bags (item number: AC-SS02). Items used for VAMS collection were purchased from Neoteryx (Torrance, California, USA).

A VAMS multi-mycotoxin extraction (42) began by transferring the VAMS tips from the plastic handles into 2 mL Eppendorf tubes, and pipetting 250 μL extraction solvent (acetonitrile/water/acetic acid, 59/40/1, *v*/*v*/*v*), containing the internal standards ^13^C_17_–AFB1 (0.125 µg/L) and ^13^C_15_ – deoxynivalenol (DON) (0.25 µg/L), ^13^C_34_–FB1 (0.25 µg/L) and ^13^C_18_–zearalenone (ZEN) (0.125 µg/L), to the sample tubes. Subsequently, samples were ultrasonicated for 30 minutes and shaken for 60 minutes at 25°C with rotation at 1,400 rpm in a Biosan TS-100 Thermo-Shaker followed by centrifugation (10 minutes at 10,000g, room temperature). The tips were discarded, and the supernatant was pipetted to an 8 mL glass tube and evaporated under nitrogen on a Turbovap LV Evaporator (Biotage, Charlotte, USA).

Afterwards, the extracts were reconstituted in 50 μL of injection solvent (methanol/water, 60/40, *v*/*v*), vortexed, centrifuged (for 10 min at 5000 g) and filtered (22 μm, PVDF, Durapore^®^,Cork, Ireland). Lastly, samples were transferred into vials before 10 μL were injected into an Acquity ultrahigh performance liquid chromatography (UPLC) system (Waters®, Manchester, UK) equipped with an Acquity HSS T3 100 × 2.1 mm UPLC column (1.8 μm particle size) and Acquity Vanguard HSS T3 10 × 2.1 mm UPLC pre-column (1.8 μm particle size), both from Waters® (Manchester, UK). Detailed instrument parameters can be found in a previous study (42).

### Statistical analysis

Data management and statistical analyses were performed using Stata (Stata Statistical Software: release 17.0; StataCorp), and a 2-sided statistical significance was considered at *p* <0.05. Descriptive statistics are presented using means ± SD or medians (range) for the continuous variables, depending on the nature of the data distribution, and frequencies (percentages) for nominal variables.

In the study sample, only exposure to OTA was found in an adequate number of newborns to assess the association with birth outcomes and infant growth. The association between OTA exposure and the study outcomes at birth and 6 months of age was evaluated using linear regression models for the continuous outcomes and linear probability models with robust variance estimation for the binary outcomes. All models were adjusted for clustering by the health center catchment areas and allocation for the prenatal and postnatal BEP interventions. Furthermore, adjusted models additionally included the covariates maternal age, primiparity, baseline BMI and hemoglobin concentration, household size, wealth index score, access to improved water and sanitation, and food security status.

We also compared OTA exposed and non-exposed groups by growth trajectories from birth to 6 months. For this purpose, we fitted mixed-effects regression models with random intercept for the individual infant and random slope for the infant’s age (months). We explored the best growth trajectory fitting the data by visual inspection of graphs and comparing model fit indices including AIC (Akaike Information Criterion) and BIC (Bayesian Information Criterion) values. Accordingly, we applied quadratic models (for the outcomes height, weight and MUAC) and restricted cubic spline model with 4 knots (for the outcome head circumference). We considered an unstructured covariance matrix for the correlation among repeated measurements within an individual. Fixed effects in the model included the main effect of OTA exposure, the main effect of age, and exposure by age interaction, which the later estimates the difference in monthly changes in the outcome between exposure and unexposed groups. Models were further adjusted for the aforementioned covariates.

In a further exploratory analysis, we evaluated potential interactions between OTA exposure and the allocation to the maternal BEP interventions on the study outcomes. For this purpose, interaction terms between OTA exposure and the prenatal and postnatal BEP interventions were specified in the models with the presence of interaction was considered at *p* <0.10.

Lastly, Cohen’s weighted kappa test was used to assess the level of agreement between mother-newborn OTA exposure status. Results were reported as percentage agreement and Cohen’s weighted kappa values. The following cut-offs were used: Kappa values ≤ 0 no agreement, 0.01–0.20 none to slight, 0.21–0.40 fair, 0.41– 0.60 moderate, 0.61–0.80 substantial, and 0.81–1.00 almost perfect agreement (43).

## Results

### Mother-newborn dyads characteristics

Birth outcomes and infant growth at 6 months were assessed on 274 and 255 newborns, respectively (**Figure 1**). Mean ± SD age of the mothers was 24.3 ± 5.63 years and 45.3% of mothers had at least a primary education (**Table 1**). The mean ± SD maternal BMI at study inclusion was 22.1 ± 3.22 kg/m^2^ with 6.93% underweight (BMI<18.5 kg/m^2^). More than two-thirds (70.3%) of the newborns were from food insecure households and 29.9% of their mothers were anemic at study enrollment during the first/early second trimester of pregnancy.

**Table 1.**
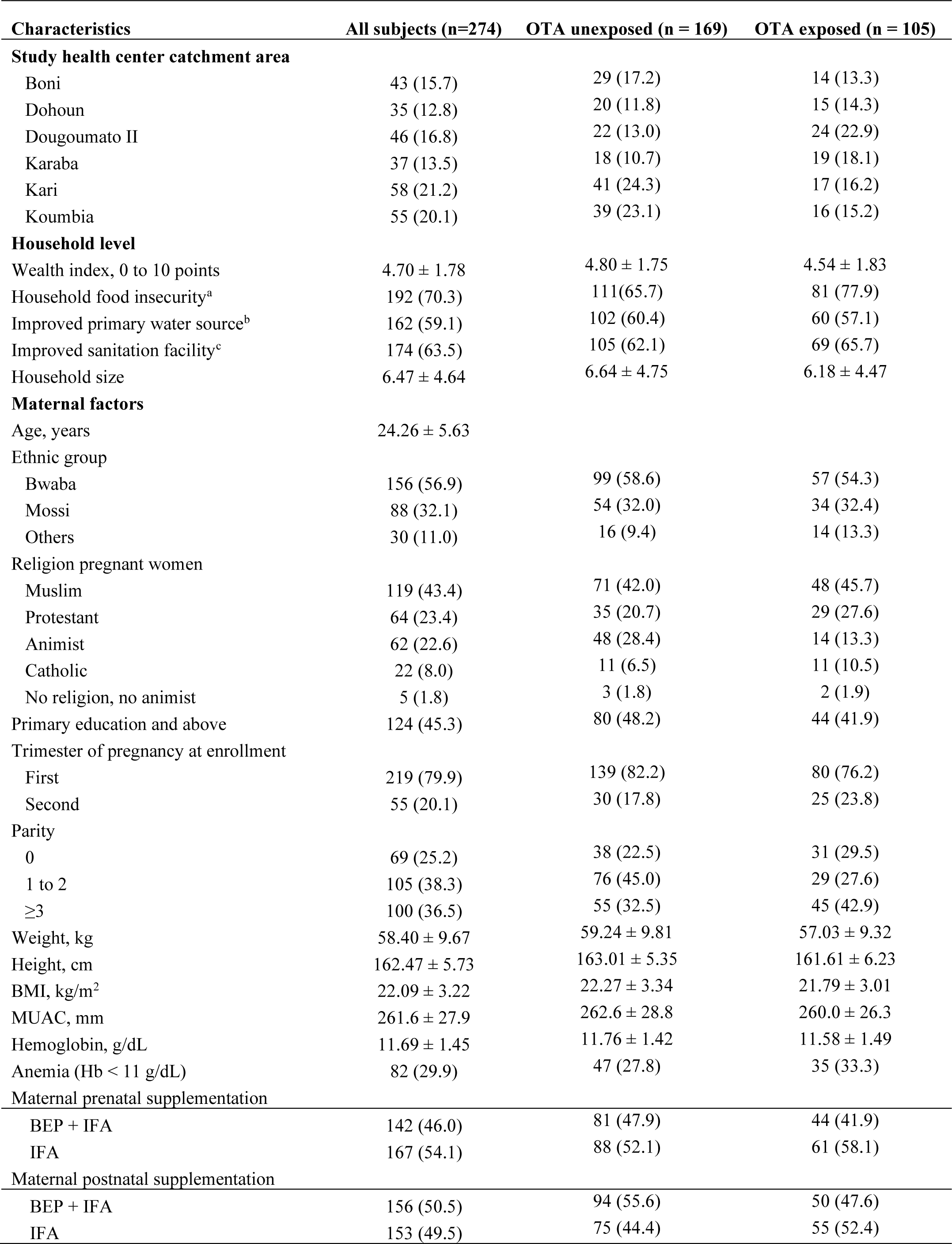
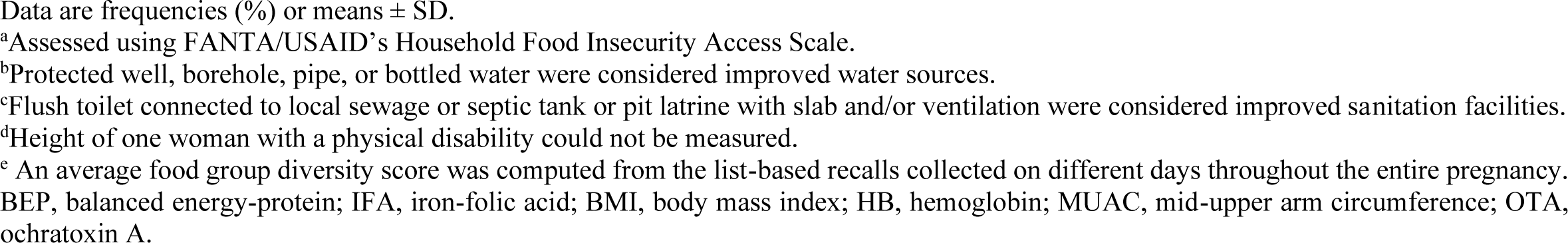
Characteristics of study participants.

| Characteristics | All subjects (n=274) | OTA unexposed (n = 169) | OTA exposed (n = 105) |
| --- | --- | --- | --- |
| **Study health center catchment area** |  |  |  |
| Boni | 43 (15.7) | 29 (17.2) | 14 (13.3) |
| Dohoun | 35 (12.8) | 20 (11.8) | 15 (14.3) |
| Dougoumato II | 46 (16.8) | 22 (13.0) | 24 (22.9) |
| Karaba | 37 (13.5) | 18 (10.7) | 19 (18.1) |
| Kari | 58 (21.2) | 41 (24.3) | 17 (16.2) |
| Koumbia | 55 (20.1) | 39 (23.1) | 16 (15.2) |
| **Household level** |  |  |  |
| Wealth index, 0 to 10 points | 4.70 ± 1.78 | 4.80 ± 1.75 | 4.54 ± 1.83 |
| Household food insecurity<sup>a</sup> | 192 (70.3) | 111 (65.7) | 81 (77.9) |
| Improved primary water source<sup>b</sup> | 162 (59.1) | 102 (60.4) | 60 (57.1) |
| Improved sanitation facility<sup>c</sup> | 174 (63.5) | 105 (62.1) | 69 (65.7) |
| Household size | 6.47 ± 4.64 | 6.64 ± 4.75 | 6.18 ± 4.47 |
| **Maternal factors** |  |  |  |
| Age, years | 24.26 ± 5.63 |  |  |
| Ethnic group |  |  |  |
| Bwaba | 156 (56.9) | 99 (58.6) | 57 (54.3) |
| Mossi | 88 (32.1) | 54 (32.0) | 34 (32.4) |
| Others | 30 (11.0) | 16 (9.4) | 14 (13.3) |
| Religion pregnant women |  |  |  |
| Muslim | 119 (43.4) | 71 (42.0) | 48 (45.7) |
| Protestant | 64 (23.4) | 35 (20.7) | 29 (27.6) |
| Animist | 62 (22.6) | 48 (28.4) | 14 (13.3) |
| Catholic | 22 (8.0) | 11 (6.5) | 11 (10.5) |
| No religion, no animist | 5 (1.8) | 3 (1.8) | 2 (1.9) |
| Primary education and above | 124 (45.3) | 80 (48.2) | 44 (41.9) |
| Trimester of pregnancy at enrollment |  |  |  |
| First | 219 (79.9) | 139 (82.2) | 80 (76.2) |
| Second | 55 (20.1) | 30 (17.8) | 25 (23.8) |
| Parity |  |  |  |
| 0 | 69 (25.2) | 38 (22.5) | 31 (29.5) |
| 1 to 2 | 105 (38.3) | 76 (45.0) | 29 (27.6) |
| ≥3 | 100 (36.5) | 55 (32.5) | 45 (42.9) |
| Weight, kg | 58.40 ± 9.67 | 59.24 ± 9.81 | 57.03 ± 9.32 |
| Height, cm | 162.47 ± 5.73 | 163.01 ± 5.35 | 161.61 ± 6.23 |
| BMI, kg/m<sup>2</sup> | 22.09 ± 3.22 | 22.27 ± 3.34 | 21.79 ± 3.01 |
| MUAC, mm | 261.6 ± 27.9 | 262.6 ± 28.8 | 260.0 ± 26.3 |
| Hemoglobin, g/dL | 11.69 ± 1.45 | 11.76 ± 1.42 | 11.58 ± 1.49 |
| Anemia (Hb < 11 g/dL) | 82 (29.9) | 47 (27.8) | 35 (33.3) |
| **Maternal prenatal supplementation** |  |  |  |
| BEP + IFA | 142 (46.0) | 81 (47.9) | 44 (41.9) |
| IFA | 167 (54.1) | 88 (52.1) | 61 (58.1) |
| **Maternal postnatal supplementation** |  |  |  |
| BEP + IFA | 156 (50.5) | 94 (55.6) | 50 (47.6) |
| IFA | 153 (49.5) | 75 (44.4) | 55 (52.4) |

Data are frequencies (%) or means ± SD.
<sup>a</sup>Assessed using FANTA/USAID's Household Food Insecurity Access Scale.
<sup>b</sup>Protected well, borehole, pipe, or bottled water were considered improved water sources.
<sup>c</sup>Flush toilet connected to local sewage or septic tank or pit latrine with slab and/or ventilation were considered improved sanitation facilities.
<sup>d</sup>Height of one woman with a physical disability could not be measured.
<sup>e</sup> An average food group diversity score was computed from the list-based recalls collected on different days throughout the entire pregnancy. BEP, balanced energy-protein; IFA, iron-folic acid; BMI, body mass index; HB, hemoglobin; MUAC, mid-upper arm circumference; OTA, ochratoxin A.

### Mycotoxins exposure and newborn and infant growth and nutritional status

The laboratory analysis indicated that, aside from OTA, almost all newborns were found to be not exposed to most mycotoxins (**Table 2**). OTA exposure was detected in 38.3% of the newborns with a median (range) concentration of <LOD (<LOD, 3.61) µg/L. The LOD for OTA as 0.09 µg/L. The UPLC-MS/MS chromatograms of OTA are shown in **Figure 3**.

**Table 2.** Newborn mycotoxin exposure at birth.

| **Mycotoxins** | **Positive samples: n (%)** | **Median (range) µg/L** |
| --- | --- | --- |
| 15-acetyldeoxynivalenol | 0.00 | <LOD (<LOD, <LOD) |
| 3-acetyldeoxynivalenol | 0.00 | <LOD (<LOD, <LOD) |
| aflatoxin B1 | 1 (0.36) | <LOD (<LOD, 0.34) |
| aflatoxin B2 | 0.00 | <LOD (<LOD, <LOD) |
| aflatoxin G1 | 6 (2.19) | <LOD (<LOD, 0.31) |
| aflatoxin G2 | 0.00 | <LOD (<LOD, <LOD) |
| aflatoxin M1 | 2 (0.73) | <LOD (<LOD, 0.7) |
| alpha-zearalenol | 0.00 | <LOD (<LOD, <LOD) |
| alternariol | 0.00 | <LOD (<LOD, <LOD) |
| alternariol monomethyl ether | 0.00 | <LOD (<LOD, <LOD) |
| beauvericin | 0.00 | <LOD (<LOD, <LOD) |
| beta- zearalenol | 0.00 | <LOD (<LOD, <LOD) |
| citrinin | 3 (1.1) | <LOD (<LOD, 18.73) |
| cyclopiazonic acid | 0.00 | <LOD (<LOD, <LOD) |
| deepoxy- deoxynivalenol | 0.00 | <LOD (<LOD, <LOD) |
| deoxynivalenol -3- glucoside | 0.00 | <LOD (<LOD, <LOD) |
| deoxynivalenol | 2 (0.73) | <LOD (<LOD, 0.74) |
| diacetoxyscirpenol | 0.00 | <LOD (<LOD, <LOD) |
| enniatin A | 0.00 | <LOD (<LOD, <LOD) |
| enniatin A1 | 0.00 | <LOD (<LOD, <LOD) |
| enniatin B | 0.00 | <LOD (<LOD, <LOD) |
| enniatin B1 | 0.00 | <LOD (<LOD, <LOD) |
| fumonisin B1 | 0.00 | <LOD (<LOD, <LOD) |
| fumonisin B2 | 0.00 | <LOD (<LOD, <LOD) |
| fumonisin B3 | 0.00 | <LOD (<LOD, <LOD) |
| fusarenone-X | 0.00 | <LOD (<LOD, <LOD) |
| neosolaniol | 0.00 | <LOD (<LOD, <LOD) |
| nivalenol | 0.00 | <LOD (<LOD, <LOD) |
| ochratoxin A | 105 (38.32) | <LOD (<LOD, 3.61) |
| ochratoxin alpha | 0.00 | <LOD (<LOD, <LOD) |
| roquefortine-C | 0.00 | <LOD (<LOD, <LOD) |
| sterigmatocystine | 0.00 | <LOD (<LOD, <LOD) |
| T-2-toxin | 0.00 | <LOD (<LOD, <LOD) |
| zearalanone | 1 (0.36) | <LOD (<LOD, 4.54) |
| zearalenone | 11 (4.01) | <LOD (<LOD, 4.94) |

LOD, limit of detection

Other mycotoxins such as AFB1, aflatoxin G1 (AFG1), aflatoxin M1 (AFM1), DON, citrinin (CIT), zearalanone (ZAN) and ZEN were detected in the range of 0.36% and 4.01% of newborns. For the remaining 26 mycotoxins analyzed, no exposure was detected through whole blood analysis.

In the unadjusted models, newborn OTA exposure was found to be negatively associated (*p* < 0.05) with birth outcomes, such as birthweight, MUAC, ponderal index and chest circumference, as well as with LAZ at the age of 6 months (**Table 3 & 4**). These associations remained significant after adjustment for relevant covariates only for birth weight (adjusted β (95% CI): −0.11 kg (−0.21, 0.00); *p* = 0.042) and ponderal index (adjusted β (95% CI): −0.62 gm/cm3 (−1.19, −0.05); *p* = 0.034). Likewise, newborns who were exposed to OTA had marginally significantly lower height growth trajectories than their counterparts without OTA exposure (adjusted β (95% CI): −0.08 (−0.15, 0.00) cm/month; *p* = 0.057) (**Figure 2**).

**Figure 2:**
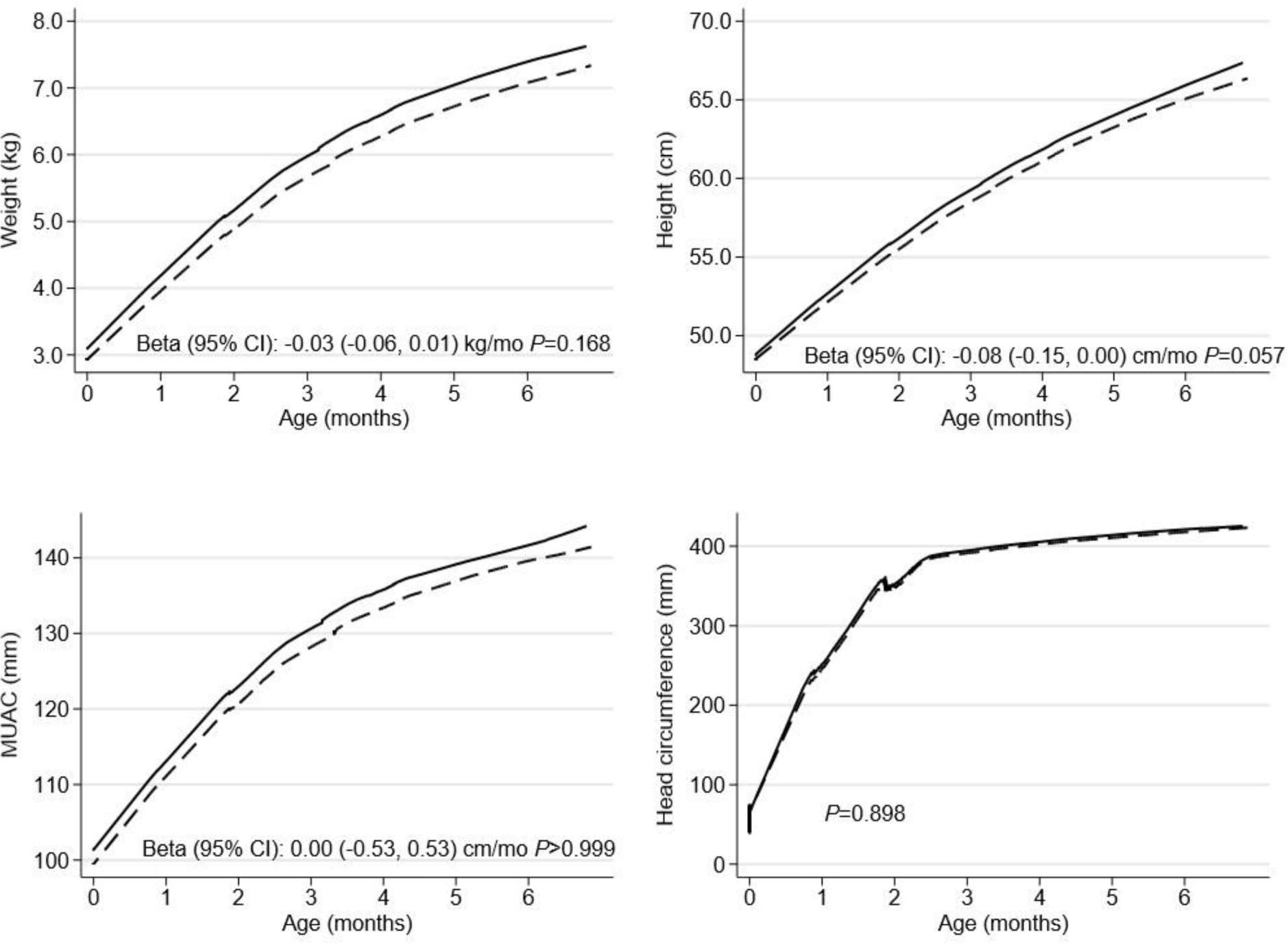
Infant growth trajectories from birth to 6 months in OTA unexposed (solid lines; *n* = 169) and exposed (dashed lines; *n* = 105) groups. Mixed-effects models with random intercept for the individual infant and random slope for the child age were fitted to compare OTA exposed and unexposed groups by growth trajectories during first 6 months postpartum. Quadratic models were used for the outcomes height, weight and MUAC and restricted cubic spline model with 4 knots for the outcome head circumference. Fixed effects in the models contained main effect of time, OTA exposure status and time by exposure interaction, which the later evaluates the difference in monthly growth trajectories between exposed and unexposed groups. Additional covariates in the models included the health center catchment areas, the prenatal and postnatal interventions allocation, maternal age, primiparity, baseline BMI and heamoglobin concentration, and household size, wealth index score, access to improved water and sanitation, and food security status. MUAC, mid-upper arm circumference; OTA, ochratoxin A.

**Figure 3.**
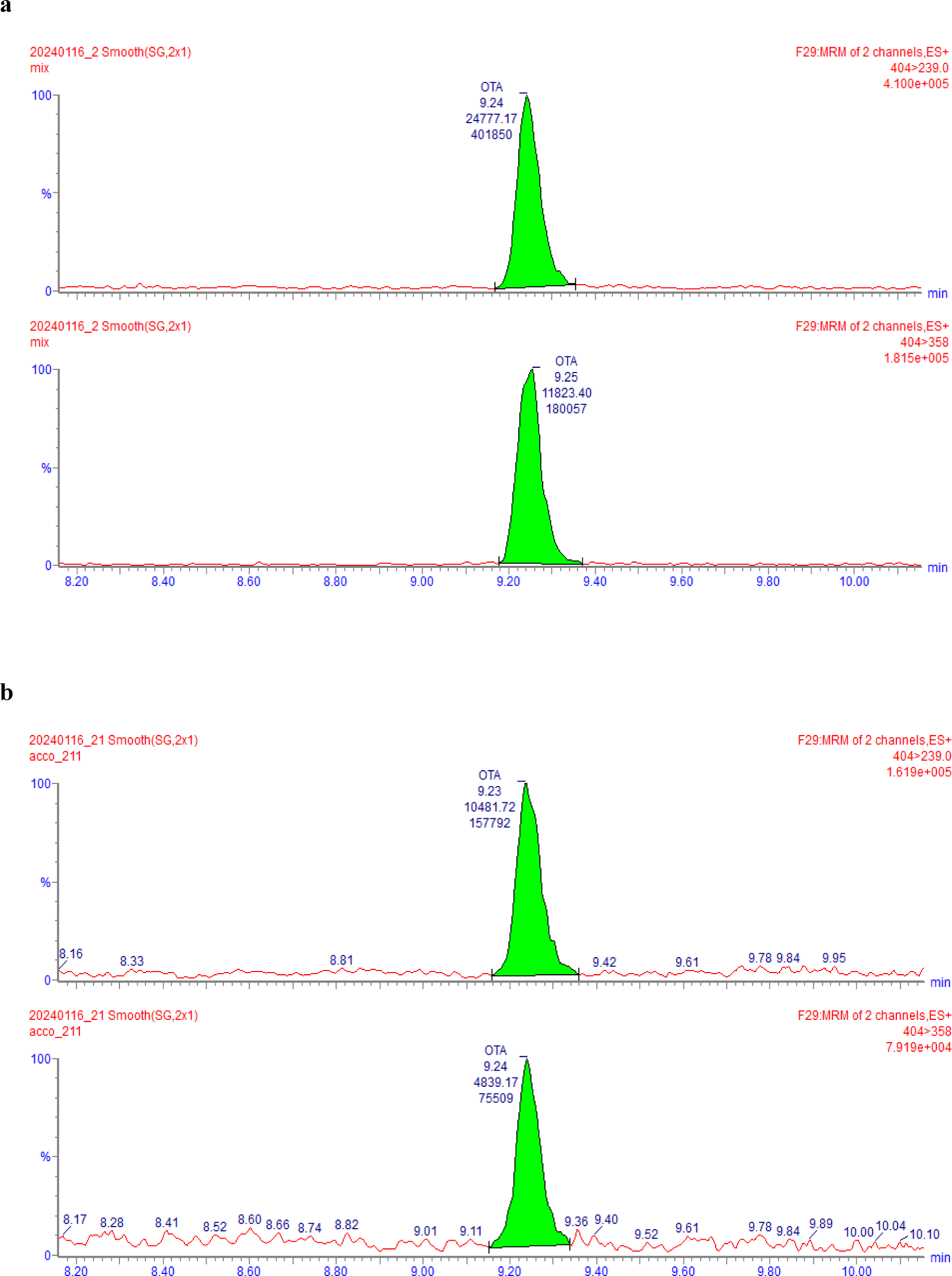
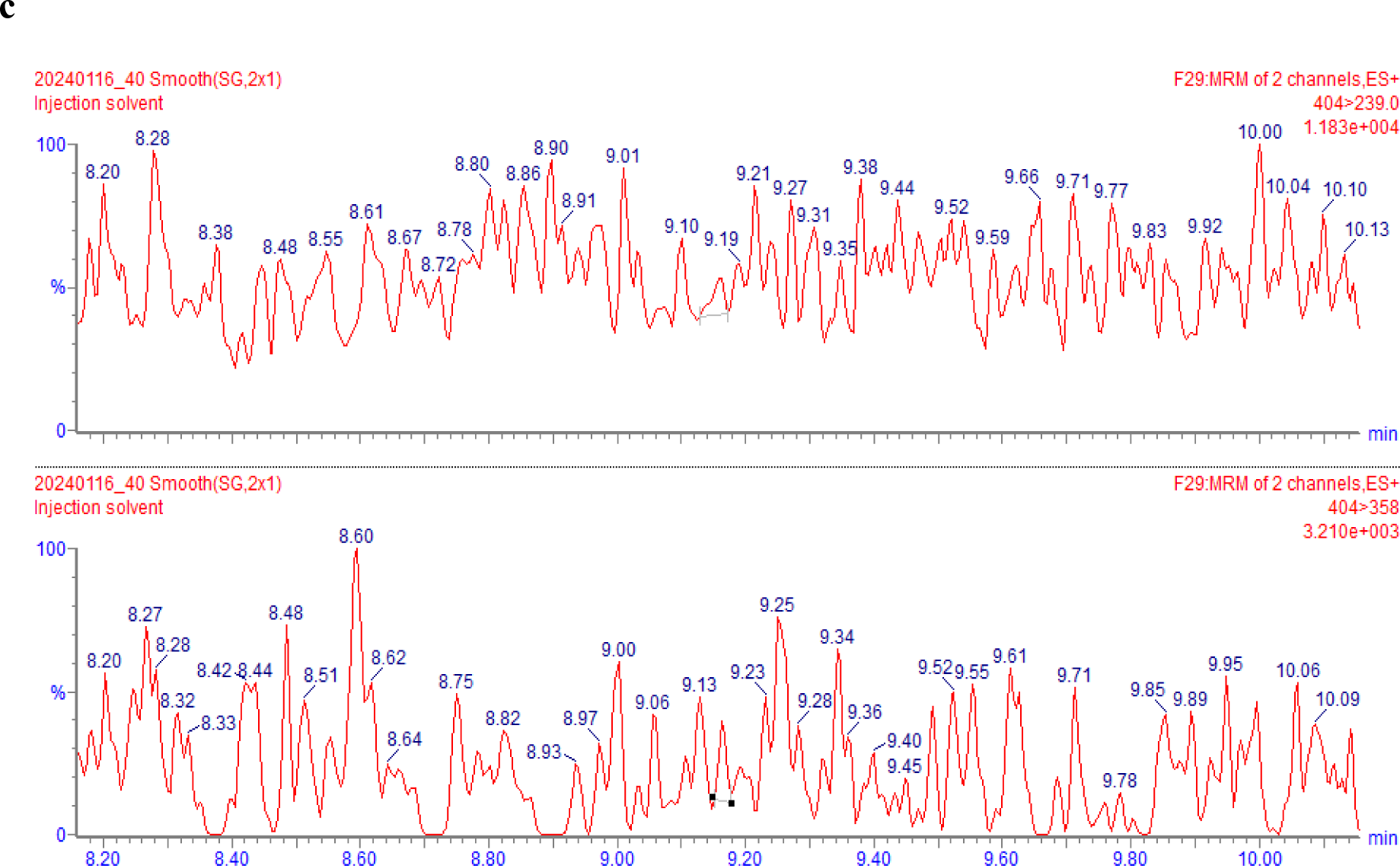
UPLC-MS/MS chromatograms of (A) OTA standard solution 2 μg/L; (B) OTA-naturally contaminated whole blood microsample (concentration 0.89 μg/L); (C) OTA-free whole blood microsample.

**Table 3:** Newborn ochratoxin A exposure and birth outcomes^1^.

| **Outcomes** | **OTA unexposed (n = 169)** | **OTA exposed (n = 105)** | **Unadjusted beta (95% CI)** | **p** | **Adjusted beta (95% CI)** | **p** |
| --- | --- | --- | --- | --- | --- | --- |
| Birthweight, kg | 3.10 ± 0.44 | 2.95 ± 0.41 | -0.15 (-0.26, -0.04) | 0.006 | -0.11 (-0.21, 0.00) | 0.042 |
| Low birthweight (<2.5 kg) | 14 (8.28) | 17 (16.19) | 6.19 (-1.69, 14.1) | 0.123 | 4.79 (3.28, 12.9) | 0.243 |
| Small-for-gestational age | 35 (20.71) | 34 (32.38) | 10.8 (-0.41, 22.0) | 0.059 | 7.80 (-3.33, 18.9) | 0.169 |
| Gestational age, week | 40.09 ± 1.20 | 39.84 ± 1.27 | -0.23 (-0.54, 0.07) | 0.141 | -0.18 (-0.49, 0.12) | 0.236 |
| Preterm delivery (<37 week) | 3 (1.78) | 2 (1.90) | 0.09 (-3.26, 3.43) | 0.959 | -0.17 (-3.85, 3.51) | 0.926 |
| Birth length, cm | 48.73 ± 2.06 | 48.49 ± 1.94 | -0.38 (-0.86, 0.11) | 0.125 | -0.18 (-0.67, 0.30) | 0.453 |
| MUAC, mm | 101.40 ± 8.24 | 99.59 ± 8.83 | -2.27 (-4.26, -0.32) | 0.023 | -1.71 (-3.6, 0.18) | 0.077 |
| Head circumference, cm | 33.43 ± 1.51 | 33.39 ± 1.38 | -0.15 (-0.49, 0.20) | 0.403 | -0.07 (-0.41, 0.27) | 0.705 |
| Ponderal index, gm/cm<sup>3</sup> | 26.68 ± 2.38 | 25.80 ± 2.48 | -0.66 (-1.24, -0.09) | 0.023 | -0.62 (-1.19, -0.05) | 0.034 |
| Chest circumference, cm | 32.13 ± 1.62 | 31.76 ± 1.63 | -0.41 (-0.82, -0.01) | 0.046 | -0.28 (-0.67, 0.11) | 0.158 |

<sup>1</sup>Values are mean ± SD or frequencies (percentages). Unadjusted and adjusted beta coefficients are estimated using linear regression models for the continuous outcomes and linear probability models with robust variance estimation for the binary outcomes. All models are adjusted for the health center catchment areas and the prenatal and postnatal interventions arms, while adjusted models additionally contained maternal age, primiparity, baseline BMI and hemoglobin concentration, and household size, wealth index score, access to improved water and sanitation, and food security status. MUAC, mid-upper arm circumference; OTA, ochratoxin A

**Table 4.** Newborn ochratoxin A exposure and infant growth and nutritional status at 6 months of age^1^.

| **Outcomes** | **OTA unexposed (n = 169)** | **OTA exposed (n = 105)** | **Unadjusted beta (95% CI)** | ***p*** | **Adjusted beta (95% CI)** | ***p*** |
| --- | --- | --- | --- | --- | --- | --- |
| Length-for-age Z-score | -0.40 ± 1.12 | -0.70 ± 1.17 | -0.33 (-0.63, -0.03) | 0.031 | -0.25 (-0.55, 0.05) | 0.105 |
| Weight-for-age Z-score | -0.34 ± 1.07 | -0.62 ± 0.99 | -0.22 (-0.50, 0.53) | 0.113 | -0.17 (-0.45, 0.10) | 0.218 |
| Weight-for-length Z-score | -0.03 ± 1.03 | -0.20 ± 1.02 | -0.08 (-0.34, 0.19) | 0.569 | -0.07 (-0.33, 0.20) | 0.607 |
| MUAC, mm | 141.11 ± 11.29 | 139.08 ± 12.50 | -2.42 (-5.17, 0.33) | 0.085 | -2.18 (-4.97, 0.61) | 0.125 |
| Head circumference, cm | 421.14 ± 15.75 | 417.84 ± 13.65 | -3.37 (-7.18, 0.44) | 0.083 | -2.90 (-6.76, 0.96) | 0.14 |
| Hemoglobin, g/dL | 10.32 ± 1.31 | 10.31 ± 1.14 | -0.21 (-0.52, 0.10) | 0.187 | -0.20 (-0.52, 0.11) | 0.204 |
| Stunting | 13 (8.23) | 10 (10.20) | 0.68 (-6.81, 8.16) | 0.859 | -0.11 (-8.23, 8.00) | 0.978 |
| Underweight | 7 (4.43) | 7 (7.22) | 2.00 (-4.53, 8.54) | 0.546 | 2.19 (-4.82, 9.20) | 0.539 |
| Wasting | 3 (1.90) | 3 (3.09) | 1.28 (-2.74, 5.30) | 0.531 | 1.53 (-2.57, 5.63) | 0.464 |
| Anemia | 108 (69.23) | 67 (70.53) | 6.74 (-4.93, 18.4) | 0.257 | 6.79 (-4.87, 18.4) | 0.252 |

<sup>1</sup>Values are mean ± SD or frequencies (percentages). Unadjusted and adjusted betas are estimated using linear regression models for the continuous outcomes and linear probability models with robust variance estimation for the binary outcomes. All models are adjusted for the health center catchment areas and the prenatal and postnatal interventions arms, while adjusted models additionally contained maternal age, primiparity, baseline BMI and hemoglobin concentration, and household size, wealth index score, access to improved water and sanitation, and food security status. MUAC, mid-upper arm circumference; OTA, ochratoxin A.

There was also a significant interaction between newborn OTA exposure status and the maternal prenatal BEP intervention on the outcome child anemia status at 6 months of age (*p*_interaction_ = 0.074) **(Supplemental Table 1**). OTA exposure was significantly associated with higher anemia prevalence among newborns of mothers who did not receive prenatal BEP supplementation (adjusted β (95% CI): 18.7% (2.00, 35.3); *p* = 0.029), while no significant association was detected among newborns whose mothers received the prenatal BEP supplementation (adjusted β (95% CI): 0.08% (−19.1, 19.3); *p* = 0.993). In the present study, there was a 63.0% agreement between mother-newborn OTA exposure status and the kappa value was classified as fair (Kappa = 0.27).

## Discussion

There is a growing concern about the potential adverse health and developmental consequences of fetal mycotoxins exposure. The present study found a high prevalence of OTA exposure (38.32%) amongst newborns at birth. Exposures to mycotoxins, such as AFG1, AFB1, AFM1, CIT, DON, ZAN and ZEN were detected in relatively fewer subjects, whereas other mycotoxins were not detected. Moreover, we found that OTA exposed newborns had significantly lower birthweight and ponderal index than their non-exposed counterparts. OTA exposed newborns also had marginally significantly lower height growth trajectories. Finally, an exploratory analysis indicated that maternal prenatal BEP supplementation may offset the effect of OTA exposure on increased anemia prevalence at 6 months of age.

This is the first study to report the level of mycotoxins exposure in newborns using whole blood microsamples. OTA is produced predominantly by some *Aspergillus*, *Monascus*, and *Penicillium* species, which frequently contaminates cereals and derived products, dried fruit, coffee, cocoa, spices, wine and cured pork products. Considering that in Burkina Faso maize is the second most cereal produced, the Burkinabé population is often exposed to OTA (44–46). A study in Sierra Leone reported OTA exposure in 25% of cord blood samples in newborns (range: 0.2-3.5 μg/L). However, the study detected a high prevalence of overall exposure to AFB1, AFM1, aflatoxicol, AFB2, AFM2, AFG1 and AFG2 (90.6%), while the present study showed limited exposure to AF except AFB1-lys which was not analyzed (11). The occurrence of OTA detected in the present study is also comparable to previous literature using adult samples. Fan *et al.* (2019) analyzed plasma samples of 260 adults in China and detected OTA in 27.7% of samples (range: 0.31-9.18 µg/L) (21), likewise in another study the OTA prevalence was 28% in serum samples from Tunisia (range: 0.12 and 11.67 µg/L) (22). On the other hand, exposure to mycotoxins other than OTA was found to be low in our study population as compared to what has been reported previously in LMICs (8,23,48,49). The variations in physicochemical properties of mycotoxins can lead to differences in their toxicokinetic profiles, which results in different excretion amounts and times. Therefore, it is conceivable that the used UPLC-MS/MS system may not detect the lowest concentrations of mycotoxin metabolites. Further research is required on mycotoxins’ stability during the processing of foodstuffs, their fate in the digestive system as well as toxicodynamic and toxicokinetic studies (50). Additionally, the knowledge of the formation process of these metabolites and the understanding of their structure and molecular mass can solve the analytical and technological challenges associated with these metabolites. Therefore, given these facts, there may be underreporting of exposure to certain mycotoxins due to their short-half lives.

The teratogenic effects of OTA have been well reported in animal studies. Reduced birth weight and craniofacial abnormalities are the most frequent reported outcomes (51). Oral administration of OTA at 5 mg/kg body weight to pregnant rats was reported to cause a reduced weight of the fetus as well as frequent hemorrhages (52–54). In addition, a single oral dose of OTA at 4 mg/kg body weight caused abortions, maternal deaths and reduction in maternal and fetal body weights (51). In the present study, results indicated a reduction of 0.11 kg in birthweight in newborns exposed to OTA compared to unexposed newborns, though previous findings that OTA exposure is associated with poor birth outcomes and infant growth have been reported inconsistently. In Uganda, AF exposure measured in mid-pregnancy was associated with LBW and smaller head circumference (55). Formerly, Jonsyn *et al*. (1995) also reported that when OTA is present in combination with AFs and their metabolites in cord blood samples, the birth weight is likely to be reduced (47). A study from Ethiopia found an association between chronic maternal AF exposure and lower fetal growth trajectories using fetal biometry from ultrasound estimates. However, the same study did not find an association of AF exposure with birth anthropometry (56). A systematic review of studies that evaluated mycotoxin exposure and infant growth also found inconsistent results (28). With the wide variation in detected mycotoxin concentrations and possible confounding factors adjusted for in these studies, it is not surprising that some found associations and others did not.

In contrast, in our previous analysis of OTA exposure during the third trimester of pregnancy in the same cohort, we did not find associations between maternal exposure and growth at birth and at 6 months of age (35). Besides this, we only found a fair level of agreement between maternal OTA exposure during the third trimester of pregnancy (50.8%) and neonatal exposure (38.3%) status (Kappa = 0.27). There was also no constant pattern in the type or quantity of AFs or OTA detected in maternal and cord blood samples in other studies. A study in Sierra Leone detected 12.5% OTA exposure in maternal serum samples versus 25.5% in cord blood samples (11); while a study in Bolivia detected OTA in 87% in the cord plasma samples versus 12.5% in the maternal plasma samples (57). A potential reason for the higher detection of OTA in the previous prenatal maternal OTA exposure analysis conducted (35) is that it was conducted at 30-34 weeks of gestation, and previous literature have reported that the level of OTA from the mother to the fetus has a higher transfer rate in the earlier stages of pregnancy compared to later (15). Furthermore, OTA distribution in the human body could also be affected by the development of placenta and physiological differences throughout pregnancy (58). Lastly, there could be seasonal variations in mycotoxin exposure status depending on food availability, and storage conditions with the maternal samples were collected between July and March (35) while the newborn samples were collected between May and October.

In the previous analysis by de Kok *et al*. (2022) maternal BEP supplementation during pregnancy and lactation were beneficial in reducing the prevalence of LBW, and improving GA, birth weight, birth length and chest circumference (59). However, iron and folic acid supplementation in the form of BEP or IFA tablets formulations did not improve anemia prevalence during pregnancy (60). Similarly, there was a high prevalence of infant anemia at 6 months of age in both intervention and control groups (61) suggesting the limited effect of maternal iron and folic acid supplementations in the form of BEP and IFA tablets formulations. On the other hand, the exploratory analysis here indicated a beneficial role of BEP in mitigating the negative effect of OTA exposure on increased infant anemia at the age of 6 months. To our knowledge, there is no other study addressing the role of nutritional supplementation on the effects of mycotoxins exposure.

The high prevalence of OTA exposure in the present study can have severe adverse consequences. After its absorption from the gastrointestinal tract, OTA binds mainly to albumin with high affinity, resulting in its long half-life (62). The OTA mechanism of action is very complex, since it is understood to be carcinogenic, hepatotoxic, immunotoxic, neurotoxic and teratogenic, based on in vitro and on animal studies (63,64). In humans, OTA exposure has been associated with the development of Tunisian Nephropathy (65), gastric and esophageal tumors (66,67), as well as testicular cancer (68). Considering the risks posed by mycotoxins in LMICs, Matumba and colleagues (2021) proposed a framework for prevention and control of mycotoxins in grains. The guideline has five pointers including: i) Sustaining plant’s strength and health; ii) Reducing toxigenic fungal population in growing plants and in storage; iii) Rapidly reducing moisture content of grains and avoid rehydration; iv) Safeguarding outer structure of seeds/grains and v) Cleaning and removing mycotoxin high risk components. The guideline also provides recommendation on how grains should be handled from production, harvesting and storage practices all the way to processing considering the factors that promote or prevent fungal contamination and subsequent production of mycotoxins in grains (69).

Some of the strengths of the present study include determination of GA using ultrasonography and the assessment of birth outcomes within 12 hours of birth, allowing the timely and robust assessment of study outcomes. The determination of mycotoxin exposure using biomarkers is also superior to the assessment in foodstuffs used in some studies (70). This is also the first application of VAMS for mycotoxin analysis in the whole blood of newborns in an LMIC setting. Considering the benefits of VAMS and the robust method developed, VAMS sampling can be considered as an alternative technique to perform a quantitative screening of mycotoxin exposure (42). The findings also provide support for future studies, using larger cohorts, with sampling using VAMS. In addition, considering the toxicokinetic profiles of the detected mycotoxins, this microsampling technique will further highlight the effect of exposure to mycotoxins on human health, enabling further associations to be made with adverse health outcomes. Lastly, as a limitation, mycotoxin exposure data from only a single time point postnatally was considered. Future studies, using repeated mycotoxins measurements, will provide an insight into the effects of mycotoxins and their physicochemical properties in relation to the timing of exposure. Moreover, further studies assessing mycotoxin exposure during the complementary feeding period in infants and young children will also provide a full picture of the burden of the problem and its effects during the critical window period in this and similar populations.

In conclusion, this study reports a high occurrence of newborn OTA exposure and an associated risk of lower birthweight, ponderal index and height growth trajectories in rural Burkina Faso. The findings emphasize the importance of nutrition-sensitive strategies to mitigate dietary OTA in the food supply, as well as adopting food safety measures in LMICs during the fetal period of development.

## Data availability request

Given the personal nature of the data, data will be made available through a data-sharing agreement. Please contact for any queries. Supporting study documents, including the study protocol and questionnaires, are publicly available on the study’s website: https://misame3.ugent.be (accessed on 07 December 2023).

## Statements and Declarations Funding

The MISAME-III main trial work was supported by the Bill & Melinda Gates Foundation (OPP1175213). The mycotoxins sampling and analysis was supported by Fonds Wetenschappelijk Onderzoek (project No G085921N). MDB was supported by the European Research Council under the European Union’s Horizon 2020 research and innovation program (grant agreement No 946192, HUMYCO). The funders had no role in the design and conduct of the study; collection, management, analysis, and interpretation of the data; and preparation, review, or approval of the manuscript.

## Conflicts of interest

The authors have no relevant financial or non-financial interests to disclose.

## Author contributions

Author Contributions: Author Conceptualization: Y.B.-M., A.A., T.D.-C., S.D.S.,CL and M.D.B.; Methodology: Y.B-M., J.E-H., S.D.S. and M.D.B.; Software: Y.B.-M., A.A., and T.D.-C.; Validation: Y.B-M., G.D.P, J.E-H., S.D.S. and M.D.B.; Formal analysis: Y.B-M and A.A.; Investigation: Y.B.-M., G.D.P, T.D.-C, J.E-H., L.O., S.D.S. and M.D.B.; Resources: Y.B.-M., T.D.-C., L.O., L.C.T., S.D.S, C.L. and M.D.B.; Data Curation: Y.B.-M., A.A., T.D.-C. and L.O.; Writing—Original Draft: Y.B.-M. and A,A.; Writing—Review and Editing: Y.B.-M., A.A., G.D.P, T.D.-C., J.E-H., L.O., L.C.T., S.D.S., C.L and M.D.B.; Supervision: T.D.-C., S.D.S, C.L., and M.D.B.; Project Administration: Y.B.-M., T.D.-C., L.O., L.C.T., C.L., S.D.S. and M.D.B.; Funding Acquisition: T.D.-C., C.L. and M.D.B.

## Ethical considerations

The study protocol was approved by the Ethical Committee of Ghent University Hospital in Belgium (B670201734334) and the Ethical Committee of Institut de Recherche en Sciences de la Santé in Burkina Faso (50-2020/CEIRES). Written informed consent was obtained from all subjects involved in the study.

## Consent to participate

Informed consent was obtained from all subjects involved in the study.

## Consent for publication

All the authors also agreed on the publication of this article.

---

**Supplemental Table 1:**
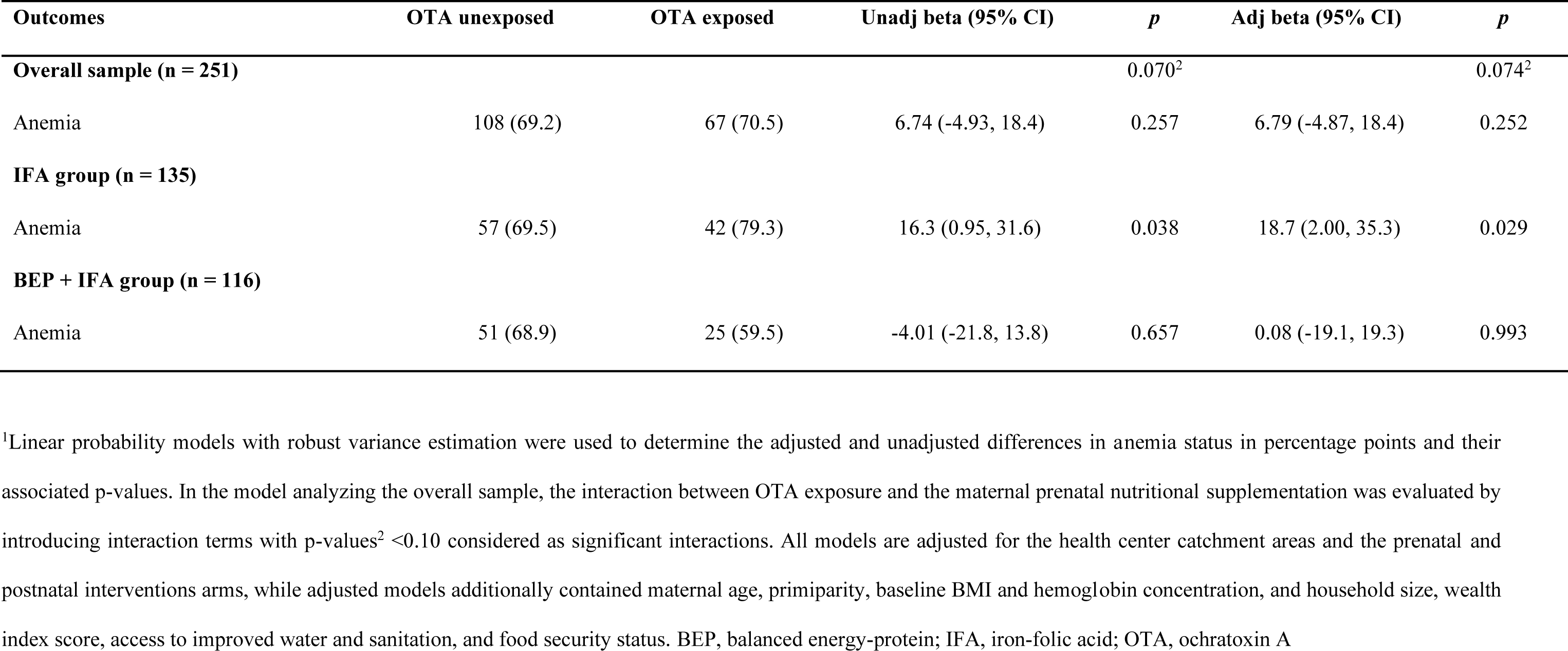
Infant anemia status at 6 months of age by OTA exposure status for the whole study sample and by maternal prenatal supplementation groups^1^.

STROBE Statement-Checklist of items that should be included in reports of cohort studies.

|  | Item No | Recommendation | Page No |
| --- | --- | --- | --- |
| Title and abstract | 1 | (a) Indicate the study's design with a commonly used term in the title or the abstract | 1<br>(Lines 1-2) |
|  |  | (b) Provide in the abstract an informative and balanced summary of what was done and what was found | 3<br>(Lines 33-55) |
| **Introduction** |  |  |  |
| Background/rationale | 2 | Explain the scientific background and rationale for the investigation being reported | 5-6<br>(Lines 59-87) |
| Objectives | 3 | State specific objectives, including any prespecified hypotheses | 6<br>(Lines 88-94) |
| **Methods** |  |  |  |
| Study design | 4 | Present key elements of study design early in the paper | 6-10 |
| Setting | 5 | Describe the setting, locations, and relevant dates, including periods of recruitment, exposure, follow-up, and data collection | 6-9 |
| Participants | 6 | (a) Give the eligibility criteria, and the sources and methods of selection of participants. Describe methods of follow-up | 6-9 |
|  |  | (b) For matched studies, give matching criteria and number of exposed and unexposed | N/A |
| Variables | 7 | Clearly define all outcomes, exposures, predictors, potential confounders, and effect modifiers. Give diagnostic criteria, if applicable | 7-9<br>(Lines 121-155) |
| Data sources/<br>measurement | 8* | For each variable of interest, give sources of data and details of methods of assessment (measurement). Describe comparability of assessment methods if there is more than one group | 6-11 |
| Bias | 9 | Describe any efforts to address potential sources of bias | N/A |
| Study size | 10 | Explain how the study size was arrived at | N/A |
| Quantitative variables | 11 | Explain how quantitative variables were handled in the analyses. If applicable, describe which groupings were chosen and why | N/A |
| Statistical methods | 12 | (a) Describe all statistical methods, including those used to control for confounding<br>(b) Describe any methods used to examine subgroups and interactions<br>(c) Explain how missing data were addressed<br>(d) If applicable, explain how loss to follow-up was addressed<br>(e) Describe any sensitivity analyses | 10-12<br>(Lines 187-221)<br>10-12<br>(Lines 187-221)<br>N/A<br>N/A<br>N/A |
| **Results** |  |  |  |
| Participants | 13* | (a) Report numbers of individuals at each stage of study—eg numbers potentially eligible, examined for eligibility, confirmed eligible, included in the study, completing follow-up, and analysed<br>(b) Give reasons for non-participation at each stage<br>(c) Consider use of a flow diagram | 12<br>(Lines 224-229)<br>N/A<br>12<br>(Lines 224-225) |
| Descriptive data | 14* | (a) Give characteristics of study participants (eg demographic, clinical, social) and information on exposures and potential confounders<br>(b) Indicate number of participants with missing data for each variable of interest<br>(c) Summarise follow-up time (eg, average and total amount) | 12<br>(Lines 224-229)<br>N/A<br>N/A |
| Outcome data | 15* | Report numbers of outcome events or summary measures over time | 12-13 |
| Main results | 16 | (a) Give unadjusted estimates and, if applicable, confounder-adjusted estimates and their precision (eg, 95% confidence interval). Make clear which confounders were adjusted for and why they were included<br>(b) Report category boundaries when continuous variables were categorized<br>(c) If relevant, consider translating estimates of relative risk into absolute risk for a meaningful time period | 12-13<br>(Lines 239-255)<br>N/A<br>N/A |
| Other analyses | 17 | Report other analyses done—eg analyses of subgroups and interactions, and sensitivity analyses | 13<br>(Lines 247-255) |
| **Discussion** |  |  |  |
| Key results | 18 | Summarise key results with reference to study objectives | 13-14<br>(Lines 257-266) |
| Limitations | 19 | Discuss limitations of the study, taking into account sources of potential bias or imprecision.<br>Discuss both direction and magnitude of any potential bias | 18<br>(Lines 367-373) |
| Interpretation | 20 | Give a cautious overall interpretation of results considering objectives, limitations, multiplicity of analyses, results from similar studies, and other relevant evidence | 13-18 |
| Generalisability | 21 | Discuss the generalisability (external validity) of the study results | 13-18 |
| **Other information** |  |  |  |
| Funding | 22 | Give the source of funding and the role of the funders for the present study and, if applicable, for the original study on which the present article is based | 19<br>(Lines 287-393) |

\*Give information separately for exposed and unexposed groups.
**Note:** An Explanation and Elaboration article discusses each checklist item and gives methodological background and published examples of transparent reporting. The STROBE checklist is best used in conjunction with this article (freely available on the Web sites of PLoS Medicine at http://www.plosmedicine.org/, Annals of Internal Medicine at http://www.annals.org/, and Epidemiology at http://www.epidem.com/). Information on the STROBE Initiative is available at http://www.strobe-statement.org.

## Notes

### Competing Interest Statement

The authors have declared no competing interest.

